# Identification of newborns at risk for autism using electronic medical records and machine learning

**DOI:** 10.1101/19008367

**Authors:** Rayees Rahman, Arad Kodesh, Stephen Z Levine, Sven Sandin, Abraham Reichenberg, Avner Schlessinger

## Abstract

**Importance:** Current approaches for early identification of individuals at high risk for autism spectrum disorder (ASD) in the general population are limited, where most ASD patients are not identified until after the age of 4. This is despite substantial evidence suggesting that early diagnosis and intervention improves developmental course and outcome.

**Objective:** Develop a machine learning (ML) method predicting the diagnosis of ASD in offspring in a general population sample, using parental electronic medical records (EMR) available before childbirth

**Design:** Prognostic study of EMR data within a single Israeli health maintenance organization, for the parents of 1,397 ASD children (ICD-9/10), and 94,741 non-ASD children born between January 1st, 1997 through December 31st, 2008. The complete EMR record of the parents was used to develop various ML models to predict the risk of having a child with ASD.

**Main outcomes and measures:** Routinely available parental sociodemographic information, medical histories and prescribed medications data until offspring’s birth were used to generate features to train various machine learning algorithms, including multivariate logistic regression, artificial neural networks, and random forest. Prediction performance was evaluated with 10-fold cross validation, by computing C statistics, sensitivity, specificity, accuracy, false positive rate, and precision (positive predictive value, PPV).

**Results:** All ML models tested had similar performance, achieving an average C statistics of 0.70, sensitivity of 28.63%, specificity of 98.62%, accuracy of 96.05%, false positive rate of 1.37%, and positive predictive value of 45.85% for predicting ASD in this dataset.

**Conclusion and relevance:** ML algorithms combined with EMR capture early life ASD risk. Such approaches may be able to enhance the ability for accurate and efficient early detection of ASD in large populations of children.

**Key points:** *Question:* Can autism risk in children be predicted using the pre-birth electronic medical record (EMR) of the parents?

*Findings:* In this population-based study that included 1,397 children with autism spectrum disorder (ASD) and 94,741 non-ASD children, we developed a machine learning classifier for predicting the likelihood of childhood diagnosis of ASD with an average C statistic of 0.70, sensitivity of 28.63%, specificity of 98.62%, accuracy of 96.05%, false positive rate of 1.37%, and positive predictive value of 45.85%.

*Meaning:* The results presented serve as a proof-of-principle of the potential utility of EMR for the identification of a large proportion of future children at a high-risk of ASD.

## Introduction

Autism Spectrum Disorder (ASD) is a neurodevelopmental disorder characterized by impairments in social communication, and restricted stereotyped behaviors^1^. Between 2000 and 2018, the number of children with ASD more than tripled, and it is now estimated that ASD affects about 1 in 59 children in the US^1^. The diagnosis of ASD typically relies on the observation of behavioral symptoms. Although these behaviors manifest at an early age (∼1 year), in the overwhelming majority of children the diagnosis is not ascertained until after the age of 4^2^. This points to an important challenge because mounting evidence indicates that early diagnosis and interventions improve the outcome for affected children^3,4^.

Existing studies have shown that ASD is highly heritable^5^. However, at present, genetic screening cannot reliably predict ASD. Despite progress in identifying rare genetic variants associated with ASD, single gene disorders only account for 3-7% of all ASD cases^6^. Thus, unlike other single-gene disorders, such as Huntington’s disease, genetic screening has limited utility in families with idiopathic ASD. In addition, due to the phenotypic heterogeneity of ASD, identification of reproducible genetic variants with significant associations to ASD incidence remains challenging^7^. Furthermore, even the strongest known ASD risk factor, a sibling with ASD, is not useful in more than 95% of ASD cases – since there is no older sibling diagnosed with ASD before the case is born. Taken together, risk-assessment models that are based on genetic information alone do not perform reliably in the context of the complex etiology of ASD.

The overwhelming majority of studies into non-genetic ASD risk factors typically consider only one exposure in isolation, e.g., paternal age^8^ or antidepressant use during pregnancy^9^. However, individual-risk factors do not provide practical predictive utility when predicting individual risk^10–12^. Furthermore, such measures of risk were often derived from studies that used traditional statistical methods to identify risk factors. Traditional statistical approaches have limited ability to handle nonlinear risk prediction and complex interactions among predictors^13^. The complex interactions between ASD in the family, mental health, medical prescriptions, and socioeconomic variables may yield greater predictive power than one factor alone.

Machine learning (ML) offers an alternative and novel analytic approach to handling complex interactions in large data, discovering hidden patterns, and generating actionable predictions in clinical settings^13–15^. Studies based on analysis of electronic medical records (EMRs) and application of ML tools have shown potential to discover complex relationships related to disease risk, from genomic studies to activities in the emergency room^13,14,16^ but, to our knowledge, has not yet been applied to address the ASD epidemic.

The aim of the current study was to test the ability of machine learning models applied to electronic medical records to predict autism spectrum disorders early in life. To address our aim, we tested the associations between an array of parental characteristics available before childbirth and the risk of ASD in offspring in a large population-based sample.

## Methods

This study was approved by the Institutional Review Board at the University of Haifa and the Helsinki Ethics Committee at Meuhedet healthcare. Those bodies waived the need for informed consent because the study data were deidentified. This study followed the Standards for Reporting of Diagnostic Accuracy (STARD) and the Transparent Reporting of a Multivariable Prediction Model for Individual Prognosis or Diagnosis (TRIPOD) reporting guidelines^17,18^.

### Data source

EMR data was obtained from a population-based case-control cohort study ascertained through a large health maintenance organization in Israel (Meuhedet). All Israeli citizens are required to purchase a medical insurance plan from one of several health maintenance organizations, which offer equivalent medical provision and fees, limiting potential selection bias in our study. Details of the ascertainment and source population, as well as, the representativeness of the cohort have been previously reported^19^. The Meuhedet cohort used in this study includes EMR data on children born in Israel from January 1, 1997, through December 31, 2007, and their parents. Children were followed up for ASD diagnosis from birth to January 26, 2015. The analytic sample consisted of 1,397 ASD cases across 1,207 father-mother pairs and, 94,741 controls across 34,912 mother-father pairs.

### Validity of ASD diagnosis

ASD diagnosis followed the International Classification of Diseases, Ninth Revision (ICD-9) and International Statistical Classification of Diseases and Related Health Problems, Tenth Revision (ICD-10). All children with suspected ASD underwent evaluation by a panel of social workers, a psychologist, and one of a trained psychiatrist, a developmental behavioral pediatrician, or a child neurologist. The final diagnosis was made by a board-certified developmental behavioral pediatrician.

### Data preparation

Parental EMR data until the child’s birth was selected. All non-drug treatments, such as medical devices, were removed along with rows containing non UTF8 formatted data. Next, a set of Anatomic Therapeutic Classification (ATC) codes were obtained from the world health organization (WHO; www.whocc.no) (2016 version). Drugs that correspond to multiple ATC classifications were combined into a single ATC code. ATC codes were mapped to drug names present in the data; multiple drug names corresponding to the same ATC or known combinations of drugs were manually annotated with unique ATC codes. Multiple prescriptions corresponding to the same ATC code within a single individual were then filtered out.

### Features set used for training

For each parent, 102 features were selected as predictors to train the different ML models. These features included prescribed medications and medical histories (e.g., number of medical contacts) (Figure 1A), as well as sociodemographic characteristics (e.g., age, socioeconomic status). Features found to be highly correlated (i.e., those exhibiting >.8 Pearson correlation coefficient) were removed. All non-categorical features were normalized using the Soft Max normalization technique (Figure 1A). Missing values were imputed using the rfImpute method of the randomForest package in the R programming language depending on ASD status^20^.

**Figure 1:**
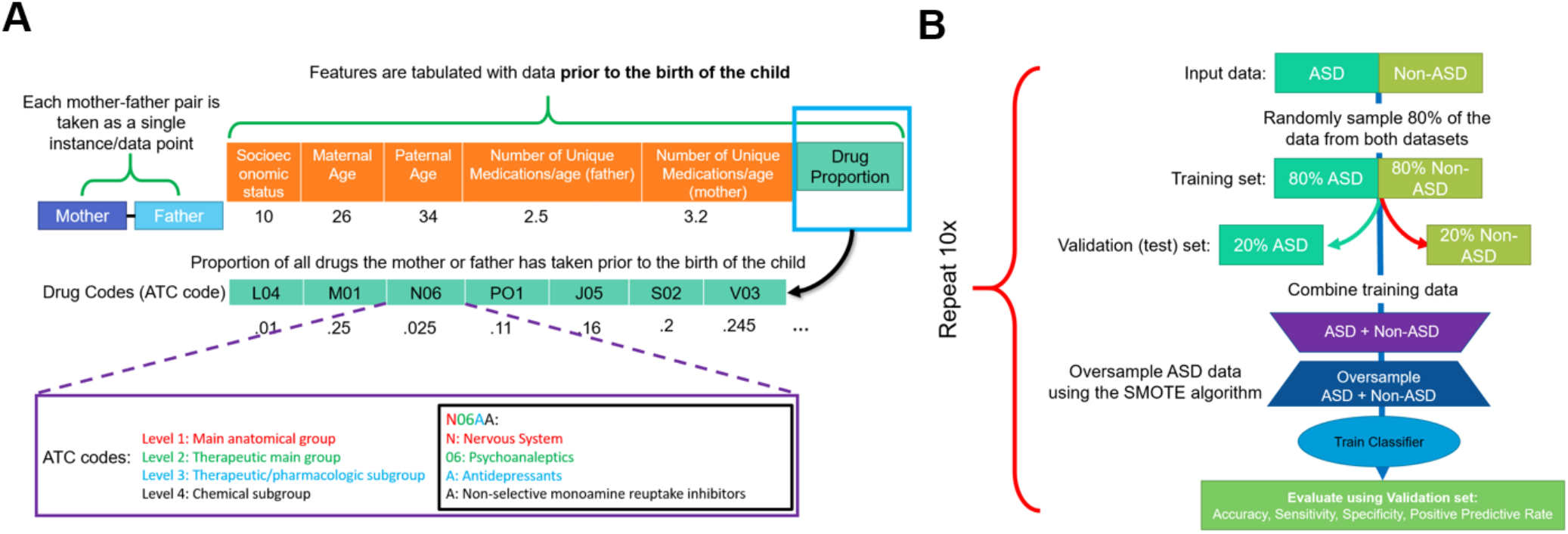
Workflow used to build machine learning model of ASD incidence. To evaluate the utility of EMR and ML for predicting the risk of having a child with ASD, we developed a comprehensive dataset. **A)** For each mother-father pair, the parental age difference, number of unique medications either parent has taken, the socioeconomic status as well as the proportion of drugs, by level 2 ATC code, taken by the parent prior to the birth of their child was used for further analysis. **B)** Workflow of performing 10-fold cross validation to evaluate model performance. First, the data was partitioned into ASD and non-ASD cases, where 80% of the data was randomly sampled as training set, and 20% was withheld as testing set. The training set was then combined and the synthetic minority oversampling technique (SMOTE) was used to generate synthetic records of ASD cases. A neural network, logistic regression, and random forest models were trained using the oversampled training data. They were then evaluated on the testing data based on sensitivity, precision, sensitivity and false positive rate. Since the testing data did not have synthetic cases, the model performance is indicative of performance of real data. This process was repeated 10 times, and average model performance was reported.

## Statistical analysis

### Machine learning (ML) models

Several statistical and ML methods were used to predict child ASD status. We applied logistic regression, artificial neural networks, and a decision tree algorithm, random forest, to predict the likelihood of offspring ASD diagnosis based on the parental features described. In particular, one advantage of a decision tree-based learning is that the importance explanatory features can be extracted after training. Random forest, an ensemble decision tree learner, was utilized for its comparable predictive performance to regression based techniques for EMR-based datasets^14,15^.

### Model evaluation and training

To evaluate each model, 10-fold cross validation was employed (Figure 1B). Eighty percent of all ASD and non-ASD mother-child pairs were sampled from the original dataset, while the remaining 20% were kept as the validation set. Due to the significant class imbalance present in the data, the Synthetic Minority Over-sampling Technique (SMOTE) was used to increase the proportion of ASD cases 5-fold with the ‘DMwR’ package in the training set, while the validation set sampling was unchanged^21^.

Oversampling under-represented data (and under-sampling overrepresented data) in training is a validated approach for developing predictive models for machine learning problems with large class imbalances. This approach has been used to improve prediction of breast cancer survivability^22^, and Alzheimer’s disease susceptibility^23^. After oversampling, the resultant dataset was used to train either the logistic regression model using the ‘glm’ method in the R programming language, a multilayer perceptron using the RSNNS package, or a random forest model implemented in the R using the ‘randomForest’ package^20^. We performed hyperparameter optimization of the machine learning algorithms using google cloud services. The final multilayer perception model used 4 hidden layers, of 5, 10, 10 and 2 nodes each, under default settings and the final random forest model used 1,000 subtrees and randomly sampled 20 features per tree. The remaining 20% of the ASD and non-ASD data was used as the validation set for the models.

The literature on prediction of health outcomes often focuses on the area under the receiver operating characteristic (ROC) curve (i.e., AUC or C statistic) rather than the full spectrum of prediction performance. However, a diagnosis of ASD is a rare outcome, and therefore relying on the C statistic alone may be biased due to either over-or underestimation^24^. For the evaluation of a clinical prediction tool, it has been recommended to report sensitivity, specificity, accuracy, false positive rate, and precision (positive predictive value, PPV) (Figure 2), to provide a more complete picture of the performance characteristics of a specific model. The validation process was repeated 10 times and the average sensitivity, specificity, accuracy, precision, false positive rate, and area under the receiver operator curve (AUC; C statistic) across all models were computed:

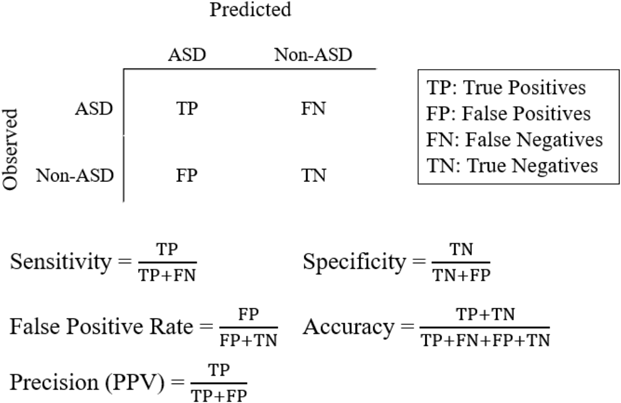

**Figure 2.**
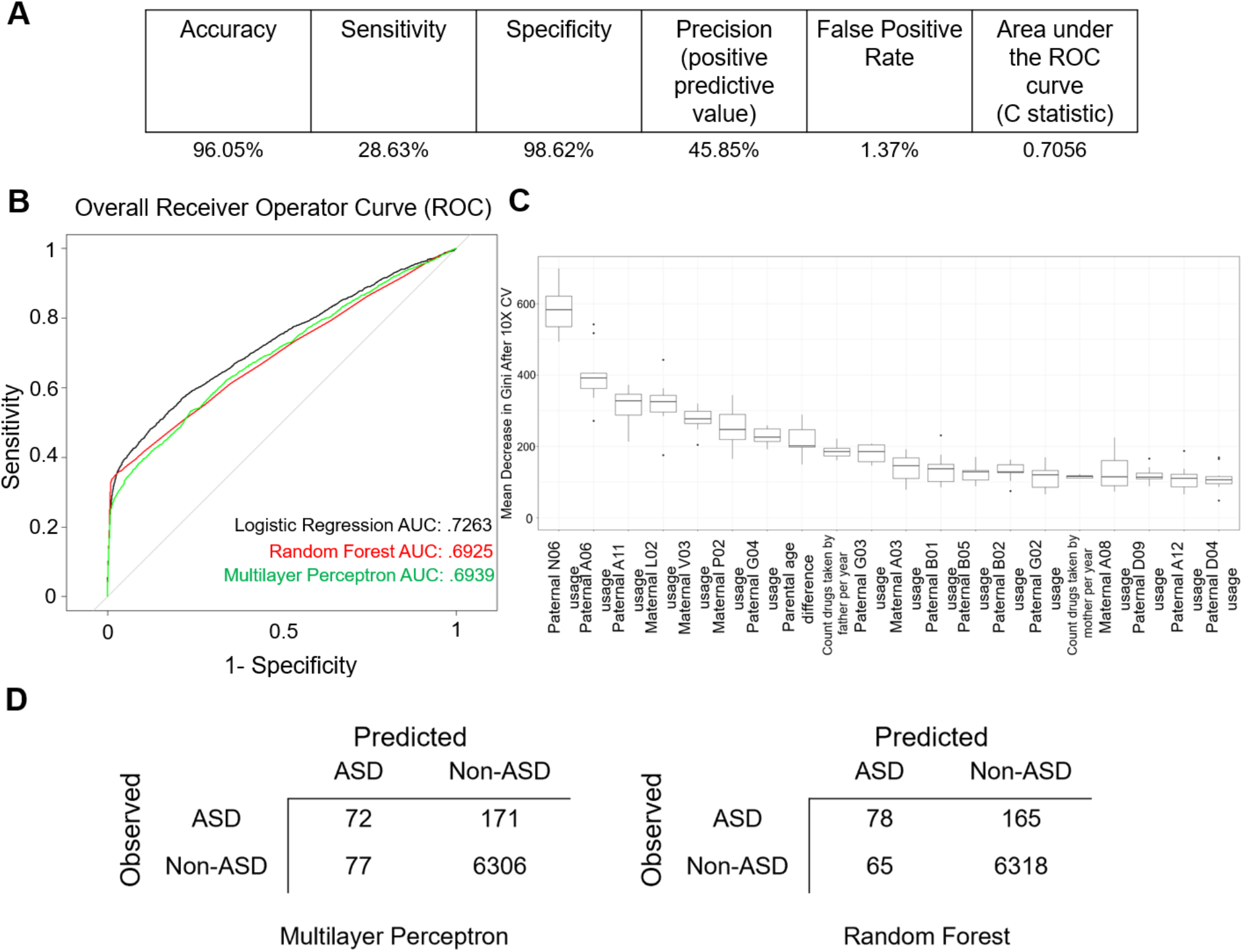
Electronic Biomarker of ASD. A balanced dataset was generated to train various algorithms to predict the probability of an ASD child from the EMR of the parents. **A**) Average validation statistics for all methods after 10-fold cross validation. **B**) Receiver operator characteristic (ROC) curves for all methods tested: logistic regression, random forest and multilayer perceptron (artificial neural network). **C**) Boxplot of importance values of each feature in the random forest model after 10-fold cross validation (10X CV). Importance of a feature is defined the mean decrease in Gini coefficient when training a model, removing the feature. Level 2 ATC codes are represented by an alphanumeric 3-letter code. D) Example confusion matrices from logistic regression and random forest models.

Notably, because oversampling only occurred during the training of the model and the validation set remained unmodified, the evaluation metrics generated are indicative of classifier accuracy for non-synthetic cases.

Gini impurity criteria was applied to determine the relative importance of individual features. The Gini impurity is the probability of an unseen case being incorrectly classified for a given decision or rule. Features with high Gini impurity (or low Gini importance) split the data into impure categories, while features that decrease Gini impurity are able to partition the data into purer classes with larger members. Thus, features with large mean decreases in Gini rank higher in importance for the model. The importance of a feature is defined as the mean decrease in the Gini impurity based on the Random Forest model.

## Results

### Prediction accuracy

After 10-fold cross validation of the testing dataset we observed that all models tested had similar performance (Figure 2), achieving an average accuracy of 96.05%, sensitivity of 28.63%, specificity of 98.62%, positive predictive value 45.85%, area under the receiver operating characteristic (ROC) curve (AUC or C statistic) of 0.70, and false positive rate of 1.37% for predicting ASD in this dataset (Figure 2A-C).

### Importance of features

The random forest model allows the identification of features with the strongest association with case classification (i.e. ASD vs. Non-ASD). Figure 2B shows the top 20 features ranked by median variable importance after 10-fold cross validation. Top features included parental age differences, and parental number of medications per year, as well as specific maternal and paternal exposure to medications. These include paternal psychoanaleptics, drugs for the treatment of blood conditions, antiparasitic medications, medications for genitourinary system and reproductive hormones, as well as nutritive supplements, maternal endocrine therapies, antheleminitics, gastrointestinal drugs and anti-obesity preparations.

### Sensitivity analysis

Machine learning (ML) models are often prone to fitting to, or ‘memorizing’, specific features or training examples, which can cause models to have poor performance for novel samples;; this is called model overfitting. Additionally, imputation of missing data can lead to a potential source of bias in our data. Since any missing features were imputed in our training data, we investigated whether our models were overfitting due to missing information. We included an additional feature labeling cases with missing parental information in order to observe the effect missing data had on predictive performance. The models were then re-generated and evaluated. The models perform similarly with or without the feature (Figure 3A), indicating that our models are unlikely to be overfitting to the lack of parental information. Finally, we also re-generated models removing features derived from either all paternal or maternal medication history (Figures 3D,3E). These models have lower C-statistic compared to those of models integrating both data sources, indicating that EMR records from either parent may explain only part of the risk of ASD in the offspring.

**Figure 3:**
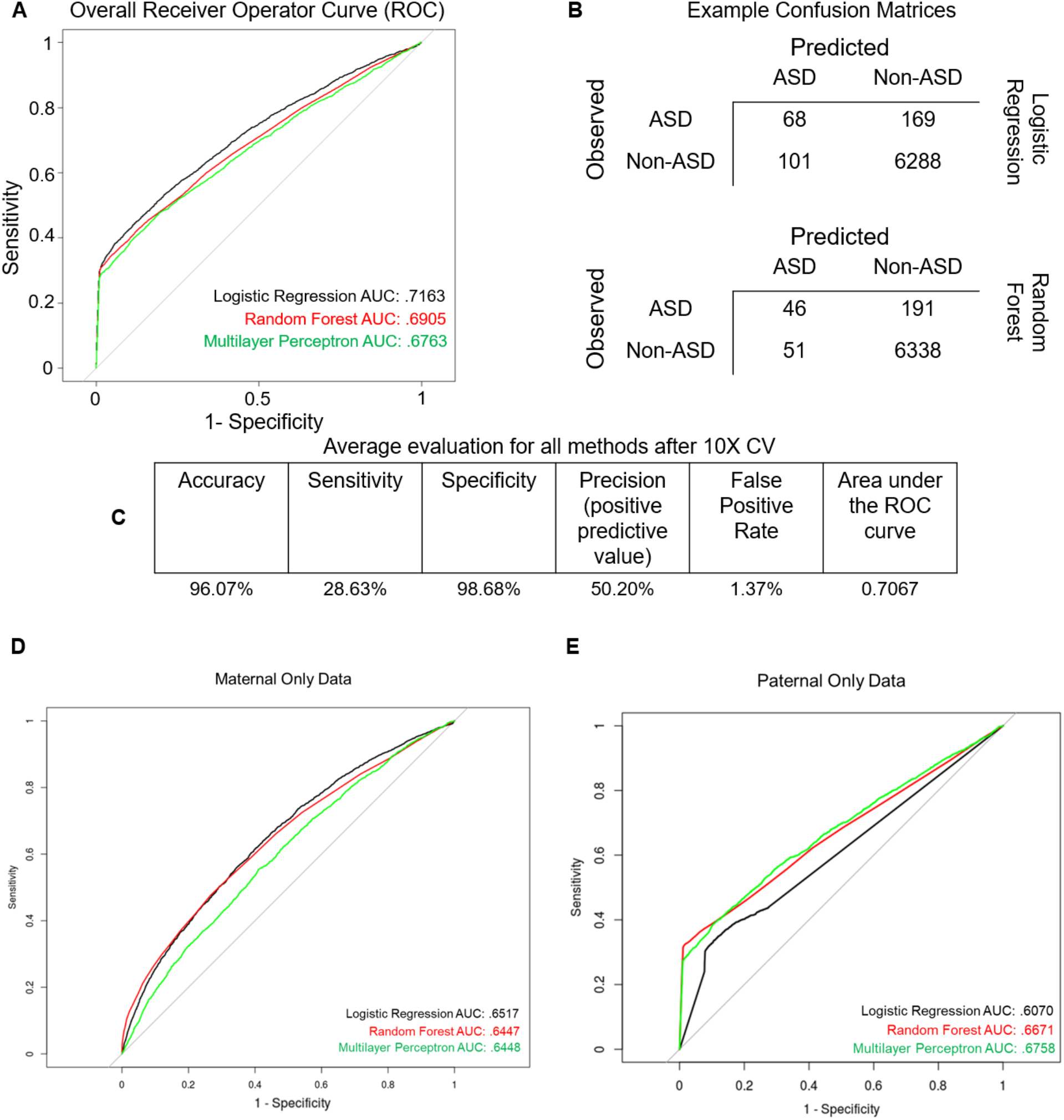
Sensitivity analysis of the generated machine learning models. **A**) Receiver operator characteristic (ROC) curves for all methods tested with ‘missing paternal information’ label included. **B**) Example confusion matrices with ‘missing paternal information’ label included. **C**) Average performance for all methods after 10-fold cross validation with ‘missing paternal information’ label included. **D**) Effect of removing all paternal medication data on model performance. **E**) Effect of removing all maternal medication data on model performance.

## Discussion

This study of population-based representative data shows that machine learning (ML) applied to EMRs could potentially identify a large proportion of future ASD cases. In the sample studied here, almost one third of ASD cases could be predicted prior to birth based on demographic and medical characteristics of their mothers and fathers. The ML models achieved high predictive performance using only routinely collected data in EMRs and without molecular genetic screening. To our knowledge, this is the first study that applied ML on EMR data to specifically predict ASD in a large population of children.

Unlike sensitivity and specificity, which are test properties, precision estimates are affected by the rate of the outcome in the population. Low rates of ASD could lead to low precision and high false-positive rate, even in tests with high sensitivity and specificity, and this therefore limit the clinical utility of a prediction algorithm. For example, screening for trisomy 21 in 20-to 30-year-old women (prevalence of approximately 1:1200)^25^, has a precision of only 1.7% with a test with sensitivity higher than 99% and specificity higher than 95%^25^. Despite the modest sensitivity in this study (28%), the specificity was very high (98%), and the precision in the study was acceptable (45%). Thus, our approach may allow high accuracy in identifying patients. To further limit the false positives in this group, additional screening and assessment may be needed.

There are several potential explanations for the gains in the prediction ability by ML. First, ML approaches possess scalability within a larger context of health information technology as they are able to extract a multitude of potential predictors from EMR. Second, ML methods are able to incorporate the high-order nonlinear interactions between features, which cannot be addressed by traditional modeling approaches (e.g. logistic regression model)^26^. Third, we applied rigorous approaches to minimize the potential of overfitting of the models.

The individual features which had the strongest association with case classification included several previously proposed sociodemographic risk factors for ASD, such as differences in parental age^8^, and parental number of medications^19^. Interestingly, there were also various medication groups that were associated with case classification. We observed a relation with psychoanaleptics, which has a mixed pattern of associations with ASD risk in previous studies^19,27,28^. We also observed a relation with nutritive supplements and reproductive hormones. Increased and decreased risk of ASD has been previously reported for medications from these groups^29,30^. Maternal metabolic conditions and disruptions in the endocrine system have been associated with ASD risk^25,26^, and contributed to case classification. Taken together, the agreement between our most useful features with previous studies analyzing ASD patients increases the confidence in our combined approach.

Notably, the associations between drugs for the treatment of blood conditions, antiparitic medications, medications for genitourinary system, anthelmintics, as well as gastrointestinal drugs that showed importance for determining ASD risk warrants further research.

Finally, we observed that apparent ASD risk can only be partially explained using either only maternal or paternal EMR data. Rather, the best predictive performance was obtained by training a model combining both sources of information. This result provides evidence that characterizing ASD incidence requires a multifaceted approach integrating maternal and paternal risk factors.

Our study has several limitations. First, the ML approaches are data driven and, therefore, depend on accurate data. While coding errors do occur, the rate of such errors in EMRs has been shown to be very low (rate less than 1%) and accuracy of the data in the Meuhedet health provider is continuously monitored for completeness and accuracy of reporting. Second, the imputation of missingness is a potential source of bias. However, the imputation by random forest is known to be a rigorous technique^31^. Third, the study lacked genetic information which could provide mechanistic interpretation of the results as well as improve prediction accuracy. Nonetheless, the objective of the present study was to develop machine learning-based prediction models that harness the readily available typical EMR data. Future studies with genetic linked data are warranted. Finally, while we show associations between several medication groups and ASD, it is important to note that this does not provide evidence that these therapeutics are causally related to ASD. Rather, parental usage of these medications may be indicative of underlying parental genetic predisposition to ASD, or the modifying role of environmental exposure on genetic predisposition.

The current study demonstrates the feasibility and potential of routinely collected EMRs data for the identification of future children at high-risk of ASD. The results also show the potential utility of data driven approaches for uncovering previously unidentified risk factors for ASD. Although certainly not causal or perfect, our results present reason for cautious optimism that recent developments in ML methodologies will be able to enhance the ability for accurate and efficient early detection of ASD in large populations of children, and allow interventions to be targeted to the small number of individuals who are at greatest risk.

## Data Availability

The data access law and ethical approvals for this study in Israel prohibits us from making individual level data publicly available. Researchers who are interested in replicating our work can apply for individual level data from the Meuhedet health care provider.

## Acknowledgment

We would like to thank Predrag Radivojac (Northeastern University) for helpful discussions and suggestions.

## Notes

### Competing Interest Statement

The authors have declared no competing interest.

### Funding Statement

No external funding was received.

### Author Declarations

All relevant ethical guidelines have been followed and any necessary IRB and/or ethics committee approvals have been obtained.

Any clinical trials involved have been registered with an ICMJE-approved registry such as ClinicalTrials.gov and the trial ID is included in the manuscript.

## References

1. Baio J. Prevalence of Autism Spectrum Disorder Among Children Aged 8 Years — Autism and Developmental Disabilities Monitoring Network, 11 Sites, United States, 2014. MMWR Surveill Summ. 2018;67. doi:10.15585/mmwr.ss6706a1

2. Kim SH, Lord C. The Behavioral Manifestations of Autism Spectrum Disorders. In: The Neuroscience of Autism Spectrum Disorders. Elsevier; 2013:25–37. doi:10.1016/B978-0-12-391924-3.00002-8

3. Christensen DL, Bilder DA, Zahorodny W, et al. Prevalence and Characteristics of Autism Spectrum Disorder Among 4-Year-Old Children in the Autism and Developmental Disabilities Monitoring Network. J Dev Behav Pediatr. 2016;37(1):1–8. doi:10.1097/DBP.0000000000000235

4. Dawson G, Rogers S, Munson J, et al. Randomized, Controlled Trial of an Intervention for Toddlers With Autism: The Early Start Denver Model. Pediatrics. 2010;125(1):e17–e23. doi:10.1542/peds.2009-0958

5. Sandin S, Lichtenstein P, Kuja-Halkola R, Hultman C, Larsson H, Reichenberg A. The Heritability of Autism Spectrum Disorder. JAMA. 2017;318(12):1182–1184. doi:10.1001/jama.2017.12141

6. Yoo H. Genetics of Autism Spectrum Disorder: Current Status and Possible Clinical Applications. Exp Neurobiol. 2015;24(4):257–272. doi:10.5607/en.2015.24.4.257

7. De Rubeis S, Buxbaum JD. Genetics and genomics of autism spectrum disorder: embracing complexity. Hum Mol Genet. 2015;24(R1):R24-31. doi:10.1093/hmg/ddv273

8. Sandin S, Schendel D, Magnusson P, et al. Autism risk associated with parental age and with increasing difference in age between the parents. Mol Psychiatry. 2016;21(5):693–700. doi:10.1038/mp.2015.70

9. Zhou X-H, Li Y-J, Ou J-J, Li Y-M. Association between maternal antidepressant use during pregnancy and autism spectrum disorder: an updated meta-analysis. Mol Autism. 2018;9. doi:10.1186/s13229-018-0207-7

10. David A et. IQ and risk for schizophrenia: a population-based cohort study. - PubMed - NCBI. https://www.ncbi.nlm.nih.gov/pubmed/?term=9403903. Accessed September 27, 2019.

11. Christensen J et. Prenatal valproate exposure and risk of autism spectrum disorders and childhood autism. - PubMed - NCBI. https://www.ncbi.nlm.nih.gov/pubmed/23613074. Accessed September 27, 2019.

12. Malmberg A et. Premorbid adjustment and personality in people with schizophrenia. - PubMed - NCBI. https://www.ncbi.nlm.nih.gov/pubmed/9715332. Accessed September 27, 2019.

13. Lo-Ciganic W-H, Huang JL, Zhang HH, et al. Evaluation of Machine-Learning Algorithms for Predicting Opioid Overdose Risk Among Medicare Beneficiaries With Opioid Prescriptions. JAMA Netw Open. 2019;2(3):e190968–e190968. doi:10.1001/jamanetworkopen.2019.0968

14. Sahni N, Simon G, Arora R. Development and Validation of Machine Learning Models for Prediction of 1-Year Mortality Utilizing Electronic Medical Record Data Available at the End of Hospitalization in Multicondition Patients: a Proof-of-Concept Study. J GEN INTERN MED. 2018;33(6):921–928. doi:10.1007/s11606-018-4316-y

15. Steele AJ, Denaxas SC, Shah AD, Hemingway H, Luscombe NM. Machine learning models in electronic health records can outperform conventional survival models for predicting patient mortality in coronary artery disease. PLOS ONE. 2018;13(8):e0202344. doi:10.1371/journal.pone.0202344

16. Taft L, Evans R, Shyu C, et al. Countering Imbalanced Datasets to Improve Adverse Drug Event Predictive Models in Labor and Delivery. J Biomed Inform. 2009;42(2):356–364. doi:10.1016/j.jbi.2008.09.001

17. pubmeddev, al BP et. Towards complete and accurate reporting of studies of diagnostic accuracy: the STARD initiative. - PubMed - NCBI. https://www.ncbi.nlm.nih.gov/pubmed/?term=12511463. Accessed September 27, 2019.

18. pubmeddev, al CG et. Transparent reporting of a multivariable prediction model for individual prognosis or diagnosis (TRIPOD): the TRIPOD statement. - PubMed - NCBI. https://www.ncbi.nlm.nih.gov/pubmed/25569120. Accessed September 27, 2019.

19. Janecka M, Kodesh A, Levine SZ, et al. Association of Autism Spectrum Disorder With Prenatal Exposure to Medication Affecting Neurotransmitter Systems. JAMA Psychiatry. October 2018. doi:10.1001/jamapsychiatry.2018.2728

20. Liaw A, Wiener M. Classification and Regression by RandomForest. Forest. 2001;23.

21. Chawla NV, Bowyer KW, Hall LO, Kegelmeyer WP. SMOTE: Synthetic Minority Over-sampling Technique. 1. 2002;16:321–357. doi:10.1613/jair.953

22. Wang K-J, Makond B, Wang K-M. An improved survivability prognosis of breast cancer by using sampling and feature selection technique to solve imbalanced patient classification data. BMC Medical Informatics and Decision Making. 2013;13(1):124. doi:10.1186/1472-6947-13-124

23. Dubey R, Zhou J, Wang Y, Thompson PM, Ye J. ANALYSIS OF SAMPLING TECHNIQUES FOR IMBALANCED DATA: AN N=648 ADNI STUDY. Neuroimage. 2014;87:220–241. doi:10.1016/j.neuroimage.2013.10.005

24. Romero-Brufau S, Huddleston JM, Escobar GJ, Liebow M. Why the C-statistic is not informative to evaluate early warning scores and what metrics to use. Crit Care. 2015;19(1). doi:10.1186/s13054-015-0999-1

25. Lutgendorf MA, Stoll KA. Why 99% may not be as good as you think it is: limitations of screening for rare diseases. J Matern Fetal Neonatal Med. 2016;29(7):1187–1189. doi:10.3109/14767058.2015.1039977

26. Kuhn M, Johnson K. Applied Predictive Modeling. New York: Springer-Verlag; 2013. https://www.springer.com/gp/book/9781461468486. Accessed August 29, 2019.

27. Kim JY, Son MJ, Son CY, et al. Environmental risk factors and biomarkers for autism spectrum disorder: an umbrella review of the evidence. The Lancet Psychiatry. 2019;6(7):590–600. doi:10.1016/S2215-0366(19)30181-6

28. Morales DR, Slattery J, Evans S, Kurz X. Antidepressant use during pregnancy and risk of autism spectrum disorder and attention deficit hyperactivity disorder: systematic review of observational studies and methodological considerations. BMC Med. 2018;16. doi:10.1186/s12916-017-0993-3

29. Li Y-M, Shen Y-D, Li Y-J, et al. Maternal dietary patterns, supplements intake and autism spectrum disorders. Medicine (Baltimore). 2018;97(52). doi:10.1097/MD.0000000000013902

30. Kosidou K, Dalman C, Widman L, et al. Maternal polycystic ovary syndrome and the risk of autism spectrum disorders in the offspring: a population-based nationwide study in Sweden. Mol Psychiatry. 2016;21(10):1441–1448. doi:10.1038/mp.2015.183

31. Tang F, Ishwaran H. Random Forest Missing Data Algorithms. Stat Anal Data Min. 2017;10(6):363–377. doi:10.1002/sam.11348

